# CASUAL SEX AMONG MSM DURING THE PERIOD OF SHELTERING IN PLACE TO PREVENT THE SPREAD OF COVID-19: RESULTS OF NATIONAL ONLINE SURVEYS IN BRAZIL AND PORTUGAL

**DOI:** 10.1101/2020.06.07.20113142

**Authors:** Alvaro Francisco Lopes de Sousa, Layze Braz de Oliveira, Artur Acelino Francisco Luz Nunes Queiroz, Hérica Emilia Felix de Carvalho, Guilherme Schneider, Emerson Lucas Silva Camargo, Telma Evangelista de Araujo, Sandra Brignol, Isabel Amélia Costa Mendes, Willi McFarland, Inês Fronteira

## Abstract

**Background:** We investigated the extent to which Brazilian and Portuguese MSM had casual sexual relations outside their homes during the period of sheltering in place for the COVID-19 pandemic.

**Methods:** An online survey was implemented in Brazil and Portugal in April, during the period of social isolation for COVID-19, with a sample of 2,361 MSMs. Recruitment was done through meeting apps and Facebook.

**Results:** Most of the sample (53.0%; considering 53.1% in Portugal and 53% in Brazil) had casual sex partners during sheltering. Factors that increased the odds of engaging in casual sex in Brazil were having group sex (aOR 2.1, 95%CI 1.3-3.4), living in a urban area (aOR 1.6, 95%CI 1.1-2.2), feeling that sheltering had high impact on daily life (aOR 3.0, 95%CI 1.1-8.3), having casual instead of steady partners (aOR 2.5, 95%CI 1.8-3.5), and not decreasing the number of partners (aOR 6.5, 95%CI 4.2-10.0). In Portugal, the odds of engaging in casual sex increased with using Facebook to find partners (aOR 4.6, 95%CI 3.0-7.2), not decreasing the number of partners (aOR 3.8, 95%CI 2.9-5.9), usually finding partners in physical venues (pre-COVID-19) (aOR 5.4, 95%CI 3.2-8.9), feeling that the isolation had high impact on daily life (aOR 3.0, 95%CI 1.3-6.7), and HIV-positive serostatus (aOR 11.7, 95%CI 4.7-29.2). Taking PrEP/Truvada to prevent COVID-19 was reported by 12.7% of MSM.

**Conclusions:** The pandemic has not stopped most of our MSM sample from finding sexual partners, with high risk sexual behaviors continuing. Public health messages for the prevention of COVID-19 need to be crafted to explicitly address sexual behavior to reduce contamination in the current moment.

**Short Summary:** The pandemic of COVID-19 did not cause changes in the sexual behavior of MSM; they continued to engage in casual sex, using drugs, having multiple partners and adopting ineffective protective measures for COVID-19.

## Background

By September 01, 2020, Brazil was still one of the most affected countries by the COVID-19 pandemic. With 121,381 deaths and 3,908,272 cases of COVID-19 officially confirmed [1] and an incidence rate (per 100,000 population) of 1837.9; Brazil ranked in the second position in the world [2]. Portugal, where the infection started spreading nearly one month before Brazil, had 58,243 confirmed cases and 1,824 deaths by COVID-19 by the same date [3], and an incidence rate (per 100,000 population) of 573.5.

Without a vaccine or effective treatment, general preventive measures for respiratory infections remain the main form of containing the spread of the virus. Minimizing the gathering and movement of people, that is, “sheltering in place” to varying degrees of strictness have been adopted by many countries, including Brazil and Portugal [4,5]. There appears to be positive effects of sheltering on reducing the speed of COVID-19 infection. However, other aspects of health, including mental and sexual health, may be suffering [6].

Social support is a known protective factor for general health, especially for LGBT communities that have historically been victims of social exclusion. Social withdrawal can trigger harmful consequences for mental and physical health. In populations where this support is more fragile, as in the case of LGBT populations, interruptions in social support can have severe negative consequences, including greater risk of exposure to COVID-19, as the disease is still not fully understood [6].

To measure the potential consequences of COVID-19 on the sexual behavior of MSM, the In_PrEP Group in Brazil and Portugal implemented an online questionnaire. In particular, the questionnaire sought to measure whether MSM were seeking casual partners during the period when shelter in place directives were in effect and which measures they were undertaking to reduce the risk of COVID-19, as well as HIV and other STIs. Brazil and Portugal were selected as they share a language and a large flow of people between these countries each year (28,210) [8], through immigration, professional and academic activities, and tourism [9].

## Methods

### Study design, population, sampling, and recruitment

This project entitled “40tena” is derived from the In_PrEP cohort study, a multicenter survey started in 2020, which carries out behavioral follow up of MSM, implemented in all 26 Brazilian states and the Federal District, and in 15 of the 18 districts of Portugal. Due to the COVID-19 pandemic, some questions were added to assess sexual behavior during the sheltering-in-place period. The research project and the presentation of these manuscripts were guided by the STROBE tool and The Checklist for Reporting Results of Internet E-Surveys (CHERRIES).

A rapid and dynamic data collection process took place in April 2020 at a time when the two countries were under sheltering directives. In both countries, the directives of the official health agencies asked the inhabitants to “shelter in place”, avoiding close contact with people outside their place of shelter as much as possible. Essential activities such as trade and some services were maintained, but with restrictions.

In total, 2,361 MSM participated in the research, out of which 1,651 (69.9%) were from Brazil and 710 (30.1%) from Portugal. The research participants were recruited by an adaptation of the Respondent Drive Sampling (RDS) method to the virtual environment, so that one participant was responsible for recruiting other individuals of the same category as his, using their networks. To meet the requirements of the method, we selected 15 MSM with different characteristics in relation to: location in the country (divided according to the regions of the two countries); race/skin color (Caucasian and non-Caucasian), age (young, adult, and older adult) and schooling level. These were the first participants and they were identified as seeds. Each participant received the link to the research website and was instructed to invite/disseminate to MSM of their social network, until obtaining a significant sample.

The seeds were identified by the two most popular [10] geolocation-based dating applications (Grindr and Hornet) by direct chat with online users, adapted to time location sampling (TLS) technique following previous methods [11-12] (Figure 01).

**Figure 01.**
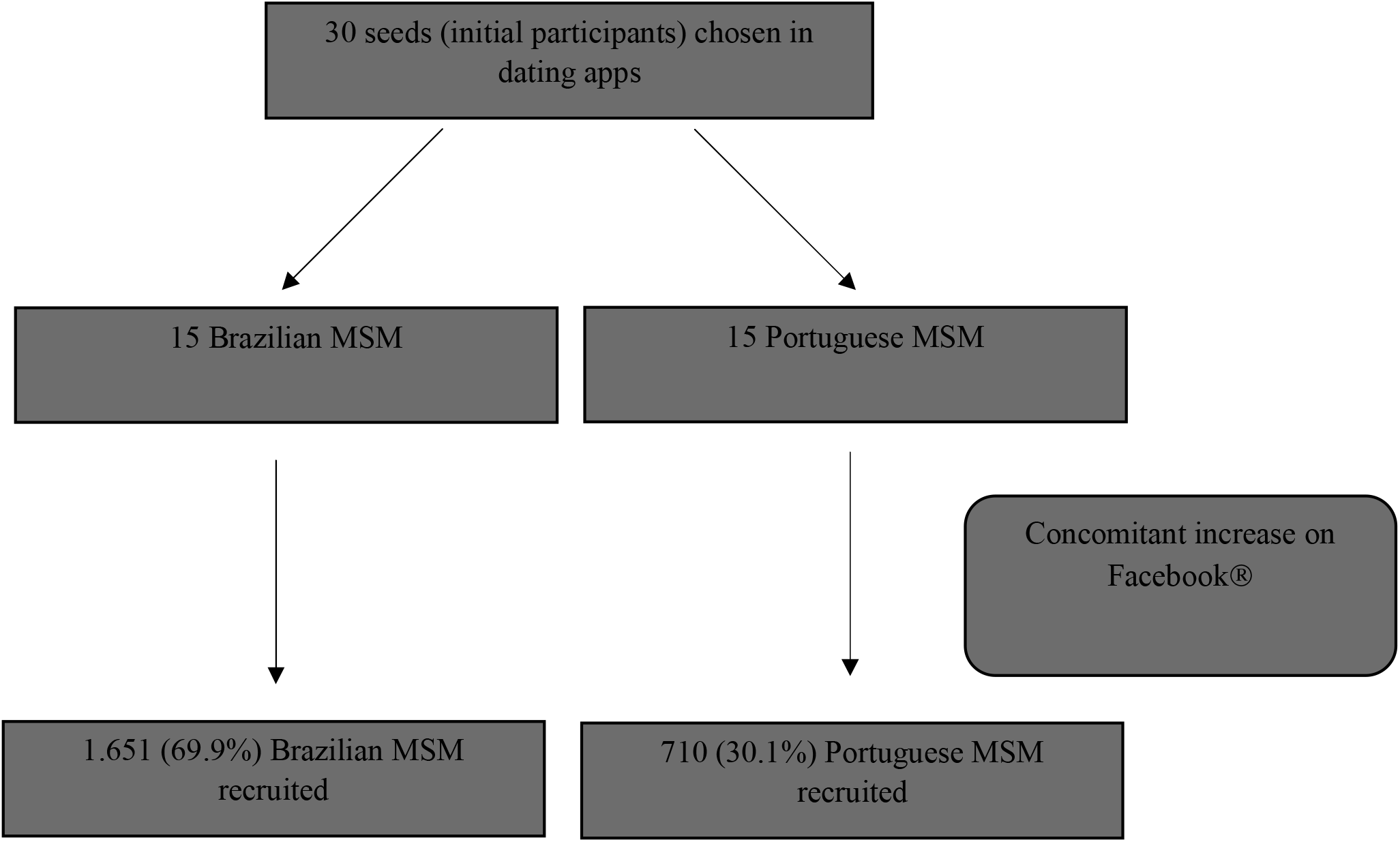
Flowchart for data collection

We also used the social network Facebook to boost the number of participants, directing to the MSM population aged between 18 and 60 years (age limit imposed by the social network), by a fixed post on the official search page (https://www.facebook.com/taafimdeque/) and it was followed by an electronic link, which provided access to the free and informed consent form and the survey questionnaire.

We included only individuals who identified themselves as men (cis or trans), aged 18 or over, and living in one of the two countries. Non-Portuguese speakers and tourists were excluded. For Facebook recruitment, the researchers used the boost feature to target MSM in both countries.

### Measures

Data were collected by Computer-Assisted Technique Interview (CASI). The data collection questionnaire was hosted on the study site and, for security reasons, it only allowed one answer by IP *(internet protocol)*, that is, only one user answered by machine (computer, cell phone…). The research questionnaire was created and validated (face-content) by a group of three experts from both countries in two versions: European Portuguese (Portugal) and Brazilian Portuguese (Brazil). It was divided into four sections with 46 questions, mostly multiple choice, some of which were mandatory for completion of the questionnaire. The questions addressed:

1. Sociodemographic information (age; gender identity; schooling; race/skin color; sexual orientation; type of current relationship; country; state; place of dwelling);
2. Social welfare and coping in the period of social distancing;
3. Sexual practices and activities during the pandemic, namely: sexual practice with a casual partner; sex with use of drugs; threesome or group sex; sex without condom use; STIs protection strategies; sex frequency and protection measures against COVID-19; search for health services;
4. Sexual practices and activities in the period prior to the pandemic: Use of PrEP and PEP; commonly used ways to search for partners; knowledge about STIs and HIV testing;

For this study, we defined sex with a casual partner or simply casual sex, such as anal sex with a new or unknown partner outside the place where they were sheltered, with the question: “Since the social distancing/sheltering was proposed in your country, have you had sex with a new or unknown partner who is outside the place where you are sheltered, or have you left your shelter to meet that partner?.”

## Analysis

Descriptive analysis was performed for key numerical and categorical variables. Bivariate and multivariable logistic regression was used to characterize associations with having casual sex.

We tested the multicollinearity between variables before moving on to the regression model. A final model was selected, using the forward conditional input method, based on retaining those variables with p<0.1 while using p<0.05 as the cut-off for significance. The best performance of the multivariate model with was considered for aspects of accuracy, sensitivity to specificity (Receiver Operating Characteristic - ROC) proving that the statistical performance developed was better than chance.

### Ethical considerations

The research project obtained ethical approval from the Universidade Nova de Lisboa and Universidade de São Paulo. Informed consent was obtained from all users online, before proceeding with the questionnaire.

## Results

A total of 2,361 MSM participated in the online surveys, including 1,651 (69.9%) from Brazil and 710 (30.1%) from Portugal (Table 1). Most participants had access to the survey from the indication by social networks partners or colleagues (72.7%), and Facebook was responsible for® the remaining 27.3%. The median age was 29 years (ranging from 18-66). Most men in both countries lived in urban areas (69.0% in Brazil, 95.4% in Portugal) and were single (69.2% in Brazil, 82.3% in Portugal). One in ten (9.9%) MSM respondents in Brazil self-reported their HIV status as positive, as did 12.1% of respondents in Portugal. In Brazil, 10.5% reported testing and 5.5% reported being diagnosed with COVID-19. In Portugal, 15.5% had tested and 1.8% were diagnosed with COVID-19.

**Table 1.**
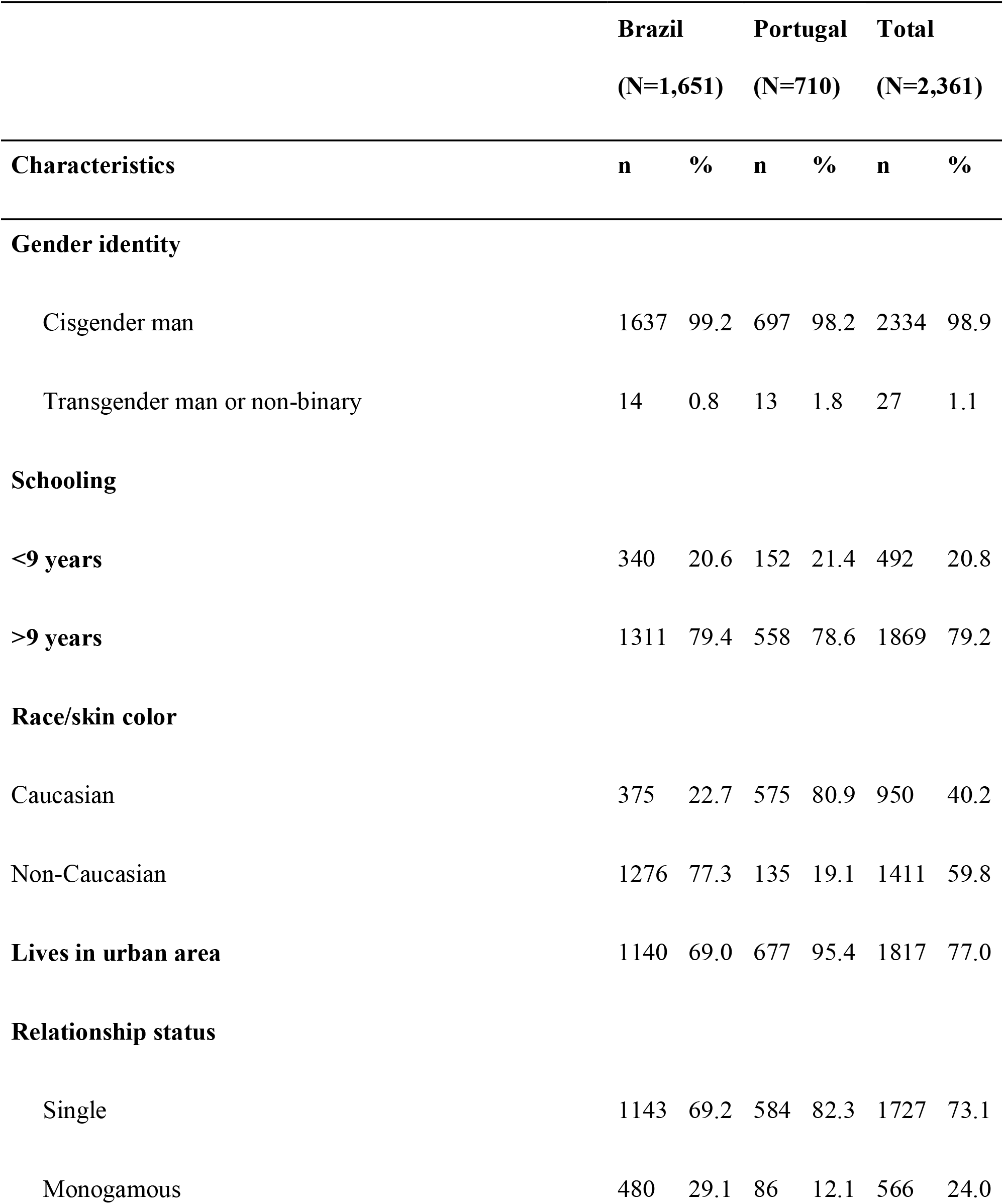
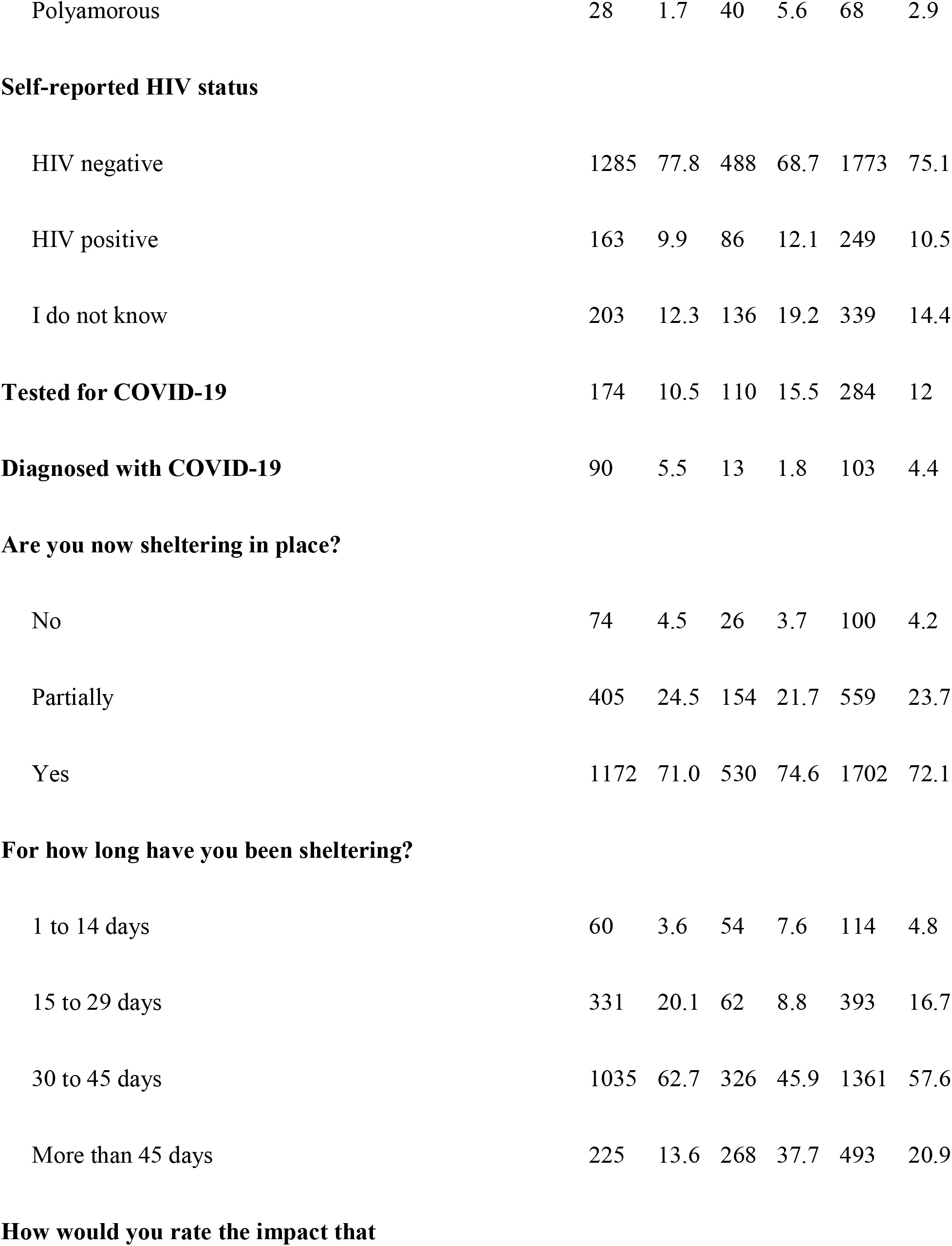
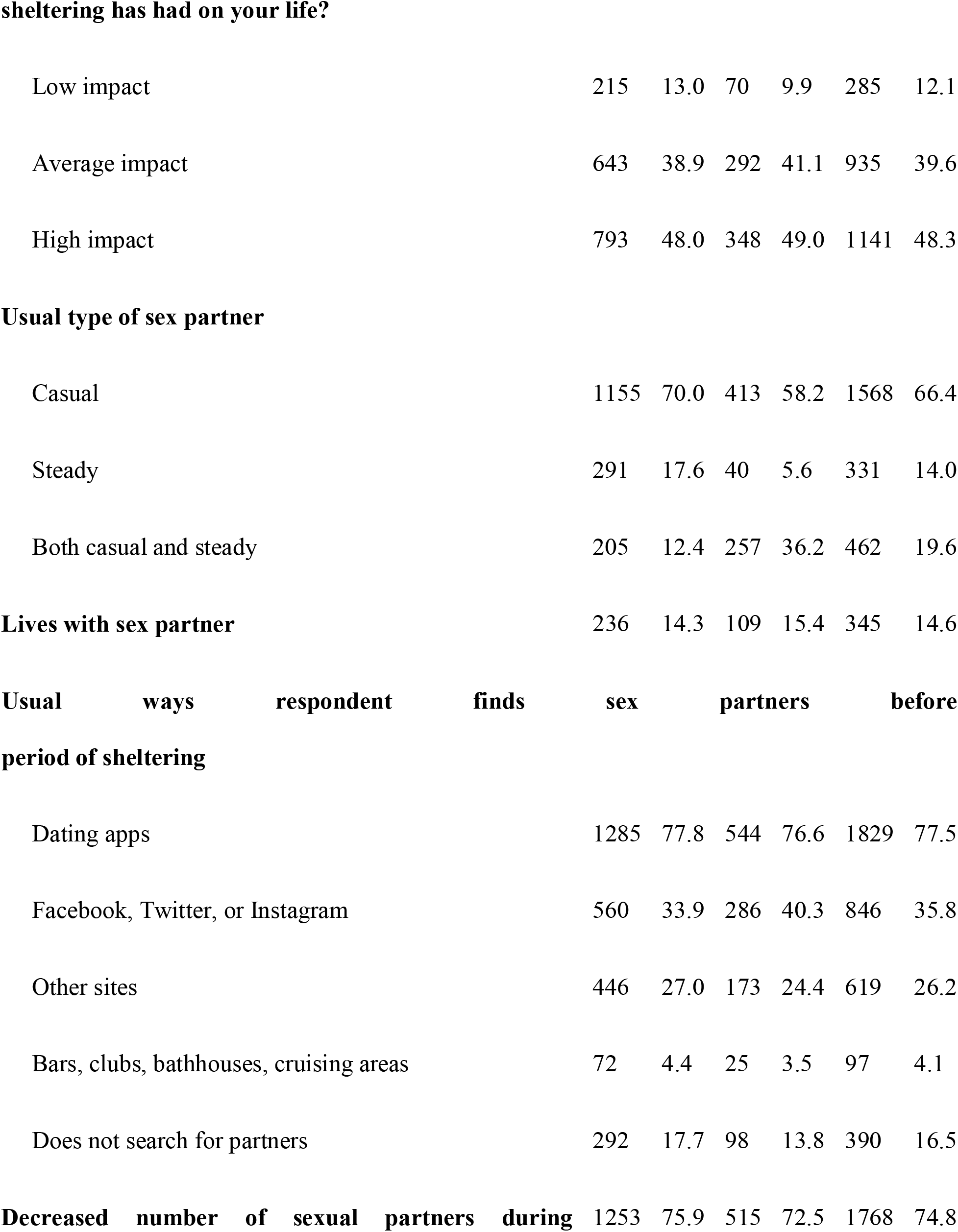
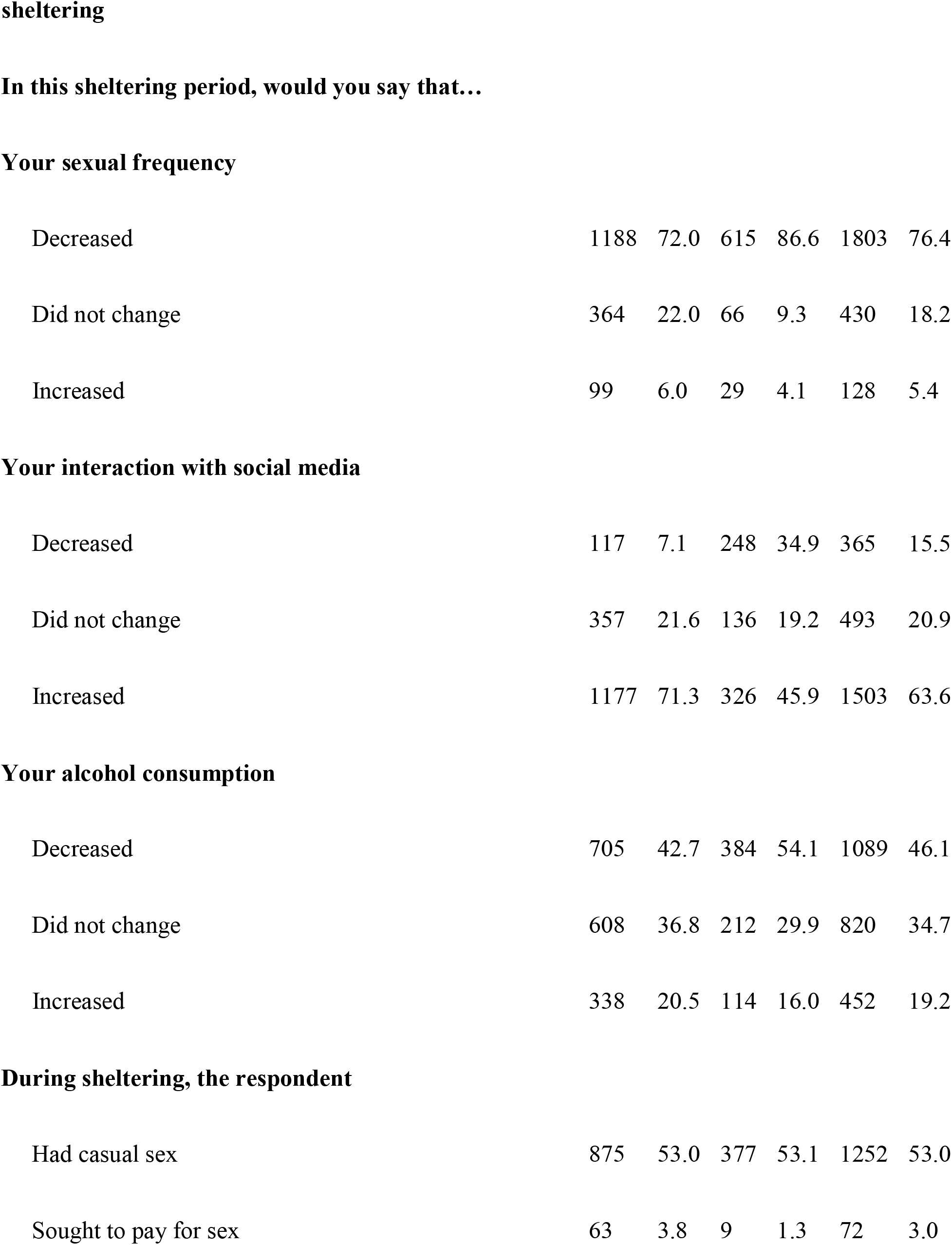
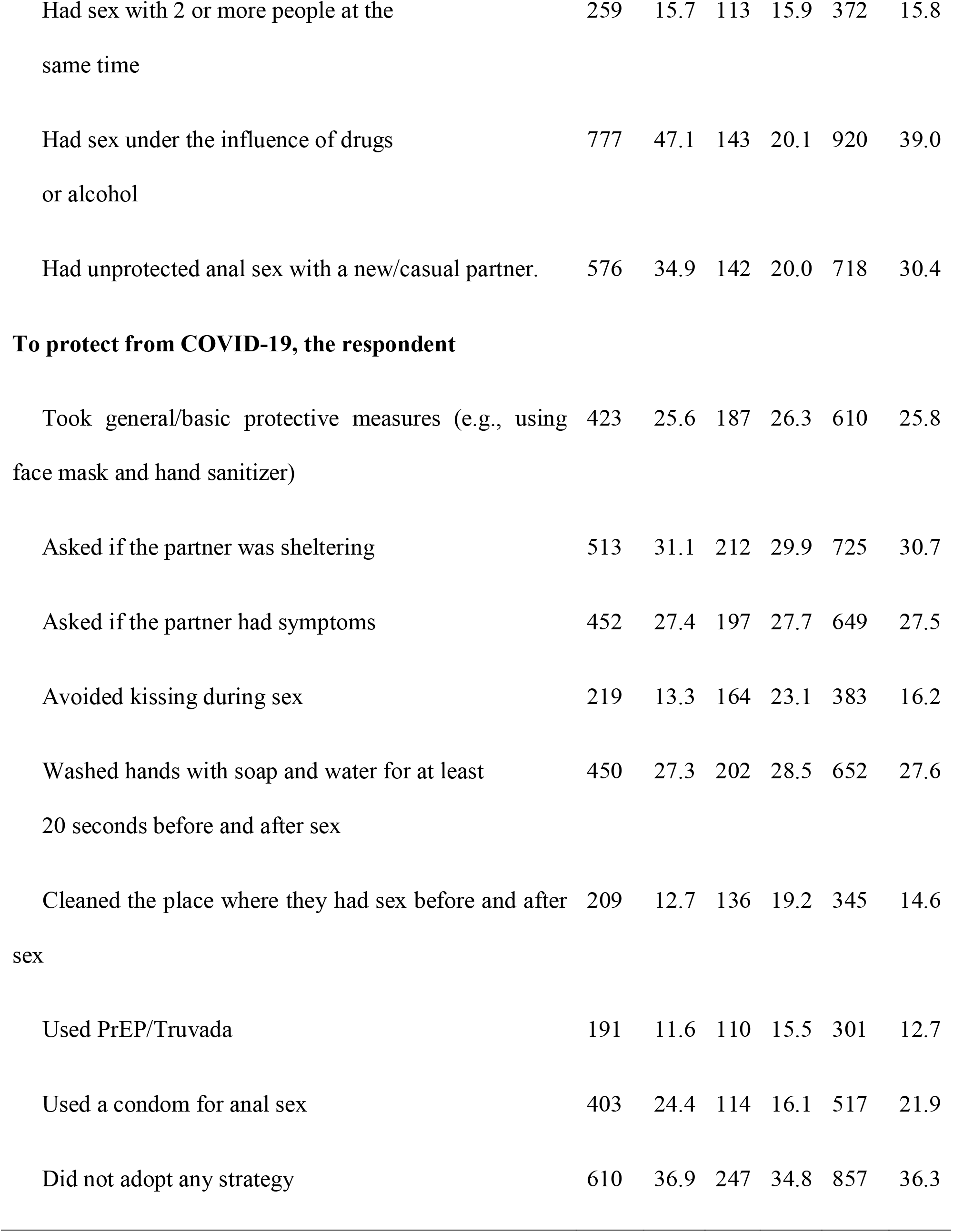
Characteristics and sexual practices during the COVID-19 shelter in place period, men who have sex with men, Brazil and Portugual, 2020.

Table 1 also describes how the COVID-19 epidemic changed the respondents’ sexual behavior. Over half of respondents (n: 1252/ 53.0%) had casual sex, considering that 47 (3.8%) engaged in paid sex, group sex (27.3%), sex under the influence of alcohol or drugs (69.8%), and unprotected sex (47.1%).

Many MSM reported behaviors that they believed would reduce the risk of COVID-19 transmission. Apart from measures taken with respect to sex, general preventive measures (e.g., use of face mask until the meeting place and hand sanitizing with hand sanitizer) (25.8%), asking if the partner was sheltering (30.7%), and asking if the partner had symptoms (27.5%) were mentioned. Other measures to reduce the spread of COVID-19 included avoiding kissing during sex (16.2%), washing hands before and after sex (27.6%), and cleaning the place where they had sex before and after sex (14.6%). Notably, among the 652 users of PrEP / Truvada in this study almost half (301; 46.1%) also referred using this medicine as a preventive measure to the transmission of COVID-19.

Table 2 presents correlates of leaving the house or having someone in their house for casual sex during the sheltering period in bivariate and multivariable logistic regression models for each country. In Brazil, the odds of engaging in casual sex increased with the variables having group sex (adjusted odds ratio [aOR] 2.1, 95% CI 1.3-3.4), living in a urban area (aOR 1.6, 95% CI 1.1-2.2), feeling that sheltering had average (aOR 2.2, 95% CI 1.5-3.2) or high impact on their daily life (aOR 3.0, 95% CI 1.1-8.3) compared to low impact, having casual partners (aOR 2.5, 95% CI 1.8-3.5), and not decreasing the number of partners during the COVID-19 epidemic (aOR 6.5, 95% CI 4.2-10.0). In Portugal, the odds of engaging in casual sex increased with the variables using Facebook to find partners (aOR 4.6, 95% CI 3.0-7.2), not decreasing the number of partners during the COVID-19 epidemic (aOR 3.8, 95% CI 2.9-5.9), usually finding partners in physical venues (pre-COVID-19) (aOR 5.4, 95% CI 3.2-8.9), feeling that the isolation had high impact on their daily life (aOR 3.0, 95% CI 1.3-6.7), and reporting HIV-positive serostatus (aOR 11.7, 95% CI 4.7-29.2).

**Table 2.**
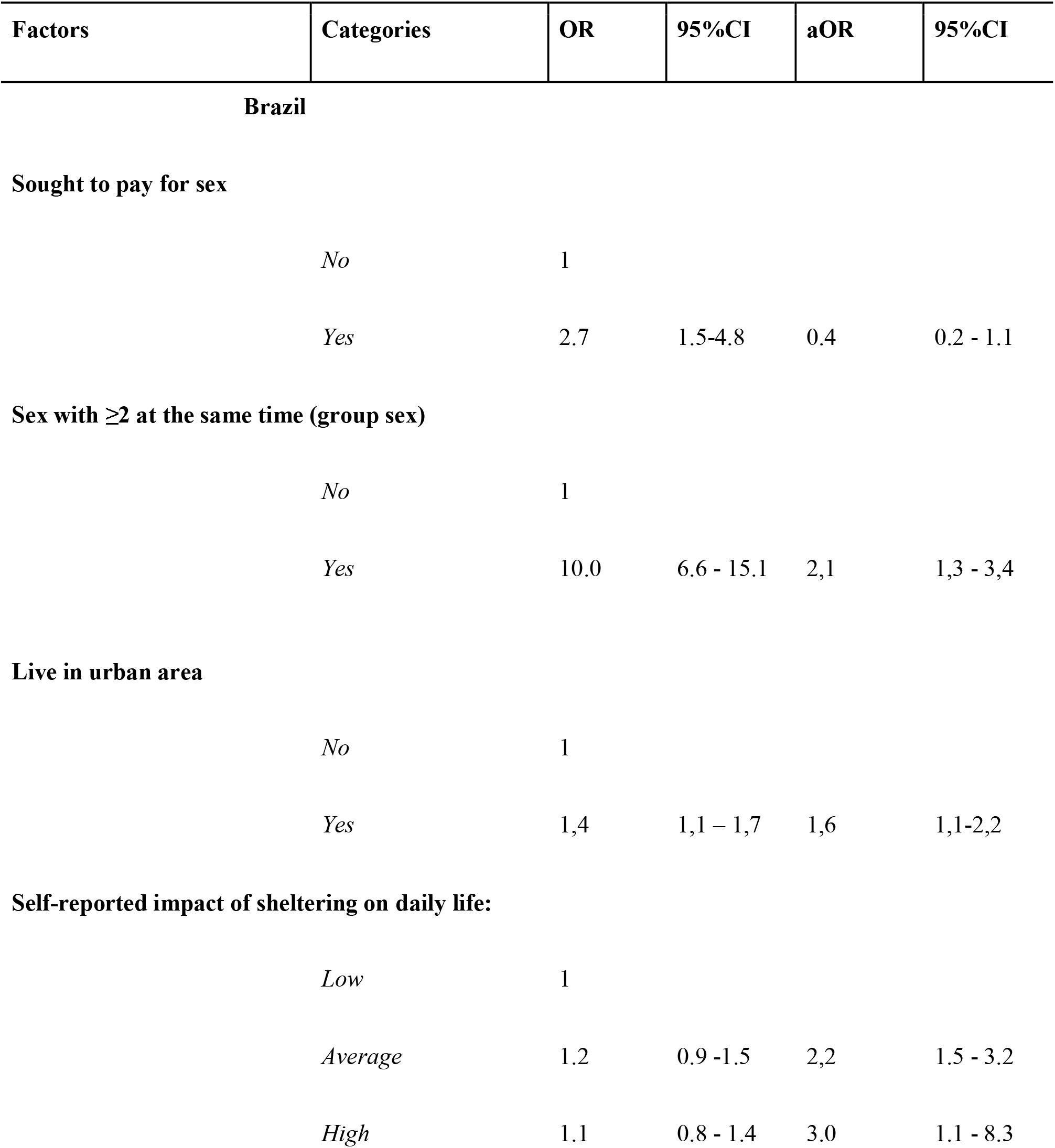
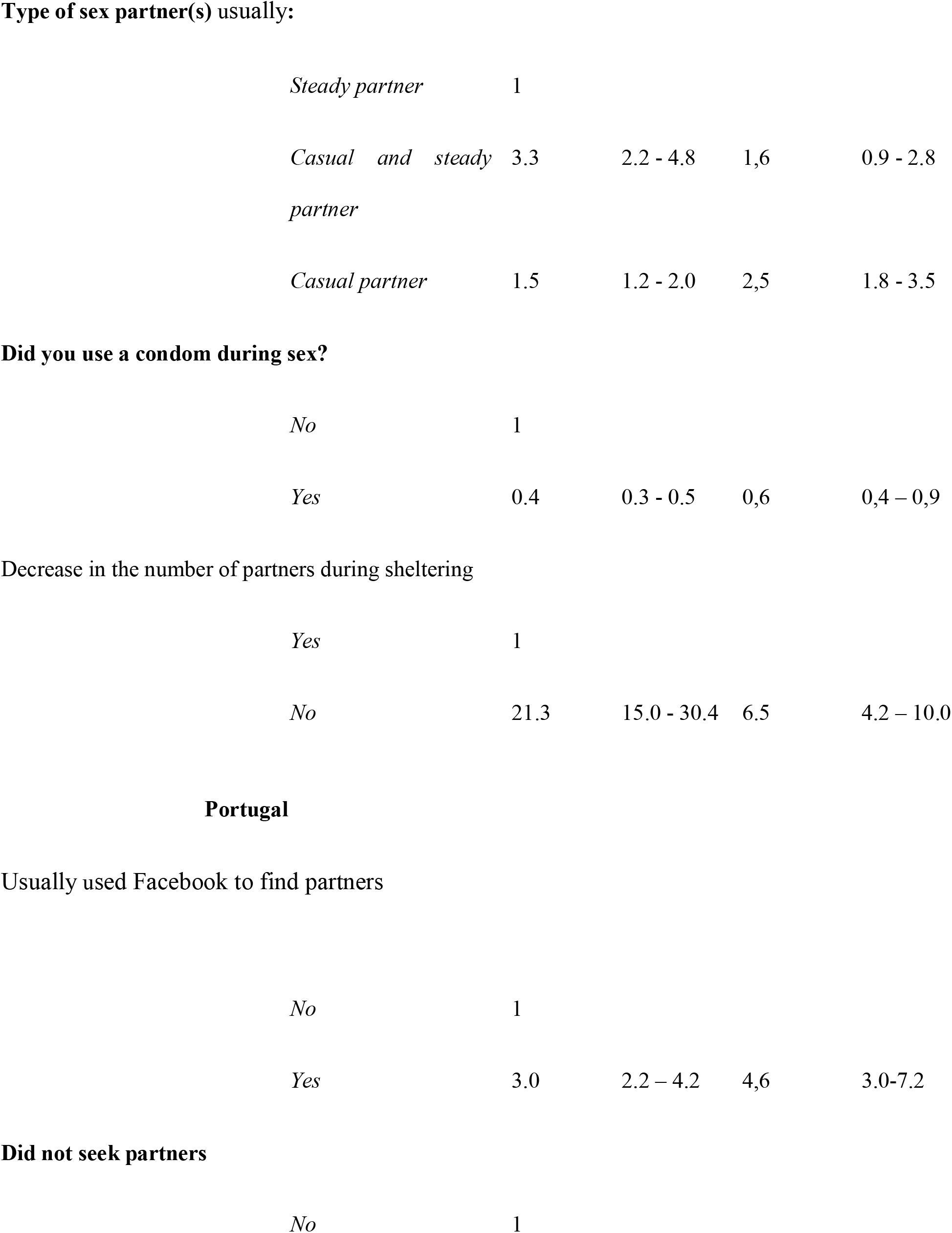
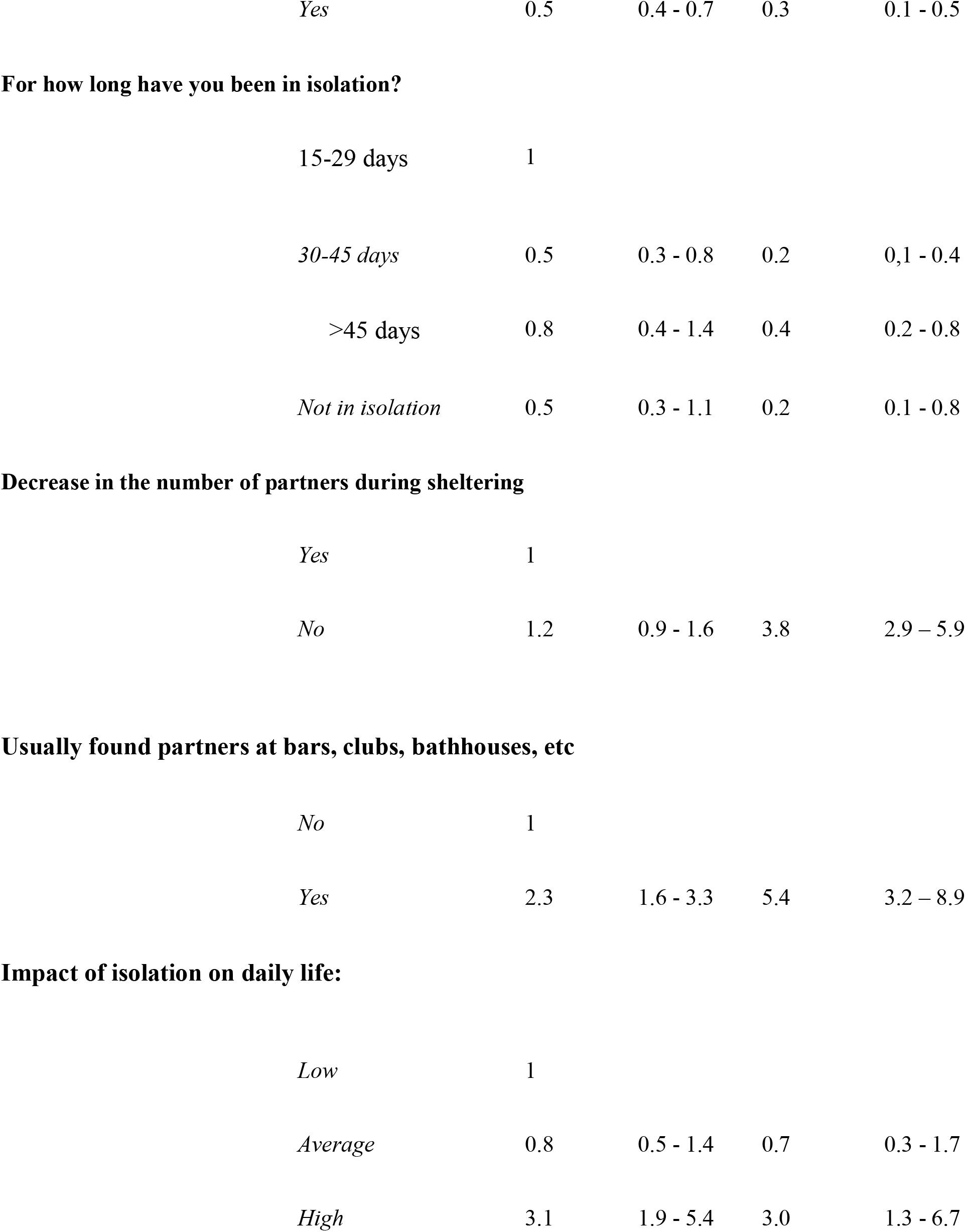
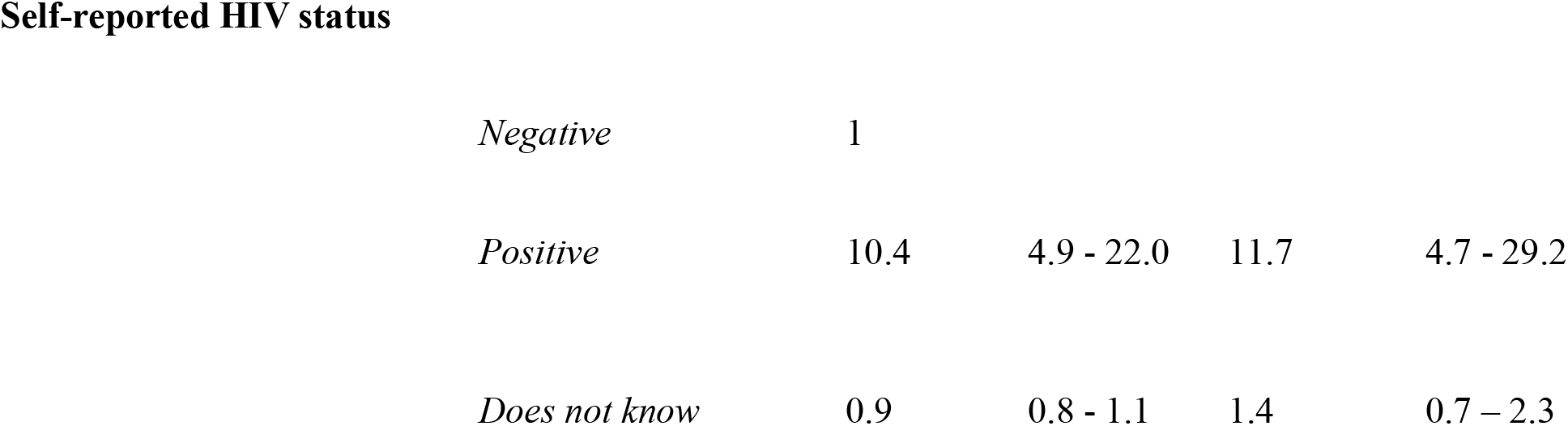
Factors associated with having casual sex during the COVID-19 shelter in place period, men who have sex with men, Brazil and Portugal, 2020.

| Factors | Categories | OR | 95%CI | aOR | 95%CI |
| --- | --- | --- | --- | --- | --- |
| **Brazil** |  |  |  |  |  |
| **Sought to pay for sex** |  |  |  |  |  |
|  | *No* | 1 |  |  |  |
|  | *Yes* | 2.7 | 1.5-4.8 | 0.4 | 0.2 - 1.1 |
| **Sex with $\geq 2$ at the same time (group sex)** |  |  |  |  |  |
|  | *No* | 1 |  |  |  |
|  | *Yes* | 10.0 | 6.6 - 15.1 | 2,1 | 1,3 - 3,4 |
| **Live in urban area** |  |  |  |  |  |
|  | *No* | 1 |  |  |  |
|  | *Yes* | 1,4 | 1,1 – 1,7 | 1,6 | 1,1-2,2 |
| **Self-reported impact of sheltering on daily life:** |  |  |  |  |  |
|  | *Low* | 1 |  |  |  |
|  | *Average* | 1.2 | 0.9 -1.5 | 2,2 | 1.5 - 3.2 |
|  | *High* | 1.1 | 0.8 - 1.4 | 3.0 | 1.1 - 8.3 |

**Type of sex partner(s) usually:**
|  |  |  |  |  |
| --- | --- | --- | --- | --- |
| *Steady partner* | 1 |  |  |  |
| *Casual and steady partner* | 3.3 | 2.2 - 4.8 | 1,6 | 0.9 - 2.8 |
| *Casual partner* | 1.5 | 1.2 - 2.0 | 2,5 | 1.8 - 3.5 |

**Did you use a condom during sex?**
|  |  |  |  |  |
| --- | --- | --- | --- | --- |
| *No* | 1 |  |  |  |
| *Yes* | 0.4 | 0.3 - 0.5 | 0,6 | 0,4 – 0,9 |

**Decrease in the number of partners during sheltering**
|  |  |  |  |  |
| --- | --- | --- | --- | --- |
| *Yes* | 1 |  |  |  |
| *No* | 21.3 | 15.0 - 30.4 | 6.5 | 4.2 – 10.0 |

**Portugal****Usually used Facebook to find partners**
|  |  |  |  |  |
| --- | --- | --- | --- | --- |
| *No* | 1 |  |  |  |
| *Yes* | 3.0 | 2.2 – 4.2 | 4,6 | 3.0-7.2 |

**Did not seek partners**
|  |  |
| --- | --- |
| *No* | 1 |

|  |  |  |  |  |
| --- | --- | --- | --- | --- |
| *Yes* | 0.5 | 0.4 - 0.7 | 0.3 | 0.1 - 0.5 |

**For how long have you been in isolation?**
|  |  |  |  |  |
| --- | --- | --- | --- | --- |
| 15-29 days | 1 |  |  |  |
| 30-45 days | 0.5 | 0.3 - 0.8 | 0.2 | 0,1 - 0.4 |
| >45 days | 0.8 | 0.4 - 1.4 | 0.4 | 0.2 - 0.8 |
| *Not in isolation* | 0.5 | 0.3 - 1.1 | 0.2 | 0.1 - 0.8 |

**Decrease in the number of partners during sheltering**
|  |  |  |  |  |
| --- | --- | --- | --- | --- |
| *Yes* | 1 |  |  |  |
| *No* | 1.2 | 0.9 - 1.6 | 3.8 | 2.9 – 5.9 |

**Usually found partners at bars, clubs, bathhouses, etc**
|  |  |  |  |  |
| --- | --- | --- | --- | --- |
| *No* | 1 |  |  |  |
| *Yes* | 2.3 | 1.6 - 3.3 | 5.4 | 3.2 – 8.9 |

**Impact of isolation on daily life:**
|  |  |  |  |  |
| --- | --- | --- | --- | --- |
| *Low* | 1 |  |  |  |
| *Average* | 0.8 | 0.5 - 1.4 | 0.7 | 0.3 - 1.7 |
| *High* | 3.1 | 1.9 - 5.4 | 3.0 | 1.3 - 6.7 |

**Self-reported HIV status**
|  |  |  |  |  |
| --- | --- | --- | --- | --- |
| *Negative* | 1 |  |  |  |
| *Positive* | 10.4 | 4.9 - 22.0 | 11.7 | 4.7 - 29.2 |
| *Does not know* | 0.9 | 0.8 - 1.1 | 1.4 | 0.7 – 2.3 |

## Discussion

Our study showed that the COVID-19 pandemic and the period of sheltering in place did not stop a considerable portion of Brazilian and Portuguese MSM from finding casual sexual partners. Nevertheless, over 95% of respondents say they adopted at least partial sheltering in place. For Brazil, this level may be higher than typically reported by local authorities for the general population (between 40% and 55%) [13]. These figures may be cause for corcern regarding overcrowding hospitals [14]. For Portugal, compliance in the general population appears to have been high enough to avert overwhelming the hospital system [15]. Although slightly over half of MSM still found casual partners outside their homes, 75% had fewer partners compared to before the COVID-19 epidemic. MSM reported other measures to reduce the risk for COVID-19 akin to other risk-reduction practices. For example, more than 25% asked if their partners were otherwise sheltering and if they had any symptoms of COVID-19. Although close contact was inherent or implied in having casual sex, many MSM reported avoiding kissing, handwashing before and after sex, and cleaning the place where they had sex before and after sex.

An unexpected finding was the use of PrEP/Truvada for COVID-19 prophylaxis. In the absence of evidence of efficacy for COVID-19 prevention, the assumption may lead to people on PrEP to neglect effective measures. A possible explanation for the adoption of this practice might be misunderstanding the discussion of potential of prophylaxis drugs for SARS-CoV2 in the popular media [16]. Some MSM may have mistaken Truvada, promoted for HIV prophylaxis, as having a similar mechanism for SARS-CoV2. Specific messaging may be needed to dispel this false connection through programs promoting PrEP for HIV.

Our study also found continuing behaviors that may place the studied group at high risk for acquiring or transmitting HIV and other STIs during the COVID-19 epidemic, at levels similar to those reported in studies [10,17] slightly prior to the pandemic period. More than one in six MSM reported group sex, implying the meeting of several people in very close contact, thus amplifying potential COVID-9 exposure [18]. Engaging in group sex was further associated with increased odds of having casual partners among the group of Brazilian MSM. Sexual encounters under the influence of drugs or alcohol, also common during the sheltering period, can decrease reasoning capacity and hinder the adoption of preventive measures for HIV/STIs and COVID-19 [19]. Unprotected sex with a new casual partner was reported by over one in three Brazilian MSM and one in five Portuguese MSM during the shelter in place period. These have been considered high rates of incidence[12].

The duration of the sheltering period, with accompanying feelings of isolation, may partly explain the high-risk sexual behaviors. Most participants had been isolated for at least 30 days, and many recognized a high impact of social isolation on their lives. This in turn may have led MSM to feel a greater need for social contact, to seek a “break” in isolation to seek partners [20], with an additional break for HIV preventive measures. This hypothesis is corroborated by the findings of the multivariable analysis, in which acknowledging high impact of the sheltering period was associated with seeking external casual sex partners in both in Brazil and in Portugal. The effect of a prolonged isolation period is particularly worrisome as Brazil moves towards becoming a COVID-19 epicenter in Latin America and the world [21].

Some studies imply that social isolation may lead to higher utilization of virtual networks to search for sexual encounters [6, 22]. Connections via the Tinder application increased 15% in the US and 25% in Italy and Spain during the COVID-19 epidemic [22]. The duration of chat activity also increased by 30% [22]. Notably, the use of Facebook as the preferable way to find partners before the pandemic was significantly associated with greater odds for Portuguese MSM having casual sex. Another hypothesis is that partnering through Facebook can provide a false sense of exposure control, as it enables sex with a friend, acquaintance or otherwise someone belonging to the same social network Yet another possible explanation for the association of increased casual partnering during COVID-19 and use of Facebook, not yet documented in the literature, may be fear of judgment (i.e., for breaking sheltering) by closer friends, which leads MSM to seek out like-minded strangers. On the other hand, social media can assist with keeping smaller social groups as well as in the adoption of practices such as virtual sex and masturbation [23, 24], which may reduce risks of transmissible infections.

Other significant associations with seeking casual sex during COVID-19 are notable. Being HIV positive also increased the odds of engaging in casual sex in Portuguese MSM. Although we recognize that even before the pandemic, studies indicated that MSM living with HIV already had high-risk sexual behavior[25,26], another hypothesis may be in the false sense of protection due to antiretrovirals for HIV currently being tested in COVID-19 patients [27]. This may be consistent with assumptions or misunderstandings about PrEP, as mentioned above.

In both Brazil and in Portugal, living in an urban area increased the odds of casual sex, likely explained by an access to greater numbers of MSM [28]. This facilitates the location and selection of partners by dating applications or other social media [12].

This study has several limitations. First, we recognize the data derived from a convenience sample in both countries. Venue-based and peer-referral mechanisms to sample and recruit are harder to logistically accomplish during the COVID-19 epidemic. The site that hosts the form is not able to inform how many subjects were reached, only the amount of access and answers.

Secondly, we did not measure variables recognized as important in hindsight, such as exact number of days in sheltering, different sexual practices, and the organization of other events, such as parties where sex may have occurred. Lastly, we did not test for COVID-19 and therefore could not fully link behaviors directly to infection. Furthermore, most questions refer to past events that may be biased by memory.

## Conclusions

We were able to identify a high frequency of casual sex among MSM, as well as associated factors that might increase exposure to SARS-CoV-2, HIV, and other STIs during a period of high COVID-19 transmission after implementation of sheltering in place. Although many strategies were adopted to minimize the exposure to SARS-CoV-2, the effectiveness of those measures is threatened by high-risk practices for both COVID-19 and HIV, including unprotected sexual intercourse and group sex. By analyzing two countries with different scenarios for control of COVID-19, we were able to demonstrate the vulnerability of MSM communities to the pandemic. Our results should be considered when making decisions about public health, since if these vulnerabilities are left unaddressed, they may hamper the response to the pandemic.

## Data Availability

Available upon request to authors.

